# Longitudinal Changes in Left Atrial Stiffness Index Improve Risk Stratification in Patients with Hypertension

**DOI:** 10.64898/2026.05.13.26353089

**Authors:** Hyue Mee Kim, Minjung Bak, Jiesuck Park, Hong-Mi Choi, Yeonyee E. Yoon, Goo-Yeong Cho, In-Chang Hwang

**Author notes:** **Address for correspondence:** In-Chang Hwang, MD, Associate Professor, Department of Internal Medicine, Seoul National University College of Medicine, Cardiovascular Center, Seoul National University Bundang Hospital, 82 Gumi-ro-173-gil, Bundang, Seongnam, Gyeonggi, 13620, South Korea.

## Abstract

**Background:** Left atrial (LA) stiffness index is a non-invasive echocardiographic parameter reflecting left ventricular filling pressure; however, its prognostic significance in hypertension remains unclear. We aimed to assess the prognostic value of the longitudinal change in LA stiffness index in patients with hypertension.

**Methods:** We analyzed 1,442 hypertensive patients from the STRATS-HHD registry who underwent echocardiography including LA and left ventricular (LV) strain at baseline and 6–18 months. Patients were categorized into four groups according to longitudinal changes in LA stiffness index: normal–normal, improved, aggravated, and persistently stiff. The primary outcome was a composite of hospitalization for heart failure (HHF) and cardiovascular death, and secondary outcomes included HHF and incident atrial fibrillation.

**Results:** Among 1,442 patients, 996 (69.1%) were classified as normal–normal, 173 (12.0%) as improved, 91 (6.3%) as aggravated, and 182 (12.6%) as persistently stiff. Over 5 years, aggravated (adjusted hazard ratio [aHR] 2.175, 95% confidence interval [CI] 1.048-4.515, P=0.037) and persistently stiff (aHR 2.935, 95% CI 1.697-5.076, P<0.001) groups were associated with a higher risk of the primary outcome, whereas the improved group showed a similar risk to the normal–normal group. Similar trends were observed for HHF and for incident atrial fibrillation. Adding LA stiffness index into a model including clinical factors and LV mass index improved risk prediction for composite outcomes.

**Conclusions:** LA stiffness index was associated with clinical outcomes in hypertension, and longitudinal changes provided additional prognostic information. Trajectory assessment may refine risk stratification.

## Introduction

Hypertension imposes a chronic hemodynamic burden on the left ventricle, leading to structural and functional remodeling, most notably left ventricular hypertrophy (LVH).^1–3^ The development of LVH is accompanied by increased left ventricular (LV) stiffness and elevated filling pressure, which are transmitted to the left atrium (LA) and lead to progressive LA remodeling.^4,5^

LA remodeling has been associated with adverse outcomes including heart failure (HF), atrial fibrillation (AF), and stroke.^6–11^ Conventional echocardiographic parameters such as LA size and LA strain have been used to assess these changes; however, they may not fully capture the interaction between LA function and LV filling pressure.^5,12,13^ The LA stiffness index, derived from LA reservoir strain (LASr) and E/e′, integrates these components and has emerged as a non-invasive marker of diastolic burden.^14^ While prior studies have linked LA stiffness index to outcomes in HF populations, its prognostic significance in hypertensive patients remains insufficiently defined.^15,16^

In hypertensive patients, antihypertensive treatment can induce regression of LVH, which has been associated with improved clinical outcomes compared with persistent or newly developed LVH.^17^ However, prior studies have primarily focused on LV remodeling, whereas the role of LA remodeling in hypertension has received relatively limited attention despite its close link to chronic diastolic burden. Accordingly, we aimed to evaluate the LA stiffness index and its longitudinal changes following antihypertensive treatment, and to determine their associations with clinical outcomes in patients with hypertension.

## Methods

### Study population

This study was based on a retrospective analysis of patients with essential hypertension enrolled in the Strain for Risk Assessment and Therapeutic Strategies in Patients With Hypertensive Heart Disease (STRATS-HHD) registry.^17–21^ We included consecutive patients who underwent transthoracic echocardiography at the time of hypertension diagnosis and had at least one follow-up examination performed after 6–18 months of antihypertensive treatment at Seoul National University Bundang Hospital and Chung-Ang University Hospital, two tertiary referral centers in Korea, between 2006 and 2021. Patients were excluded if they had: (1) specific cardiomyopathies, including dilated, hypertrophic, restrictive, ischemic, or stress-induced cardiomyopathy, as well as Fabry disease or mitochondrial disorders; (2) moderate or greater valvular heart disease; (3) end-stage renal disease; (4) prior cardiac surgery; or (5) other cardiovascular conditions that could contribute to left ventricular hypertrophy, including secondary hypertension. Among the initially eligible 1,600 patients, 158 were excluded due to unavailable values required for the calculation of LA stiffness index, including E/e′ and LA reservoir strain. The final study population comprised 1,442 patients (**Figure 1**).

**Figure 1.**
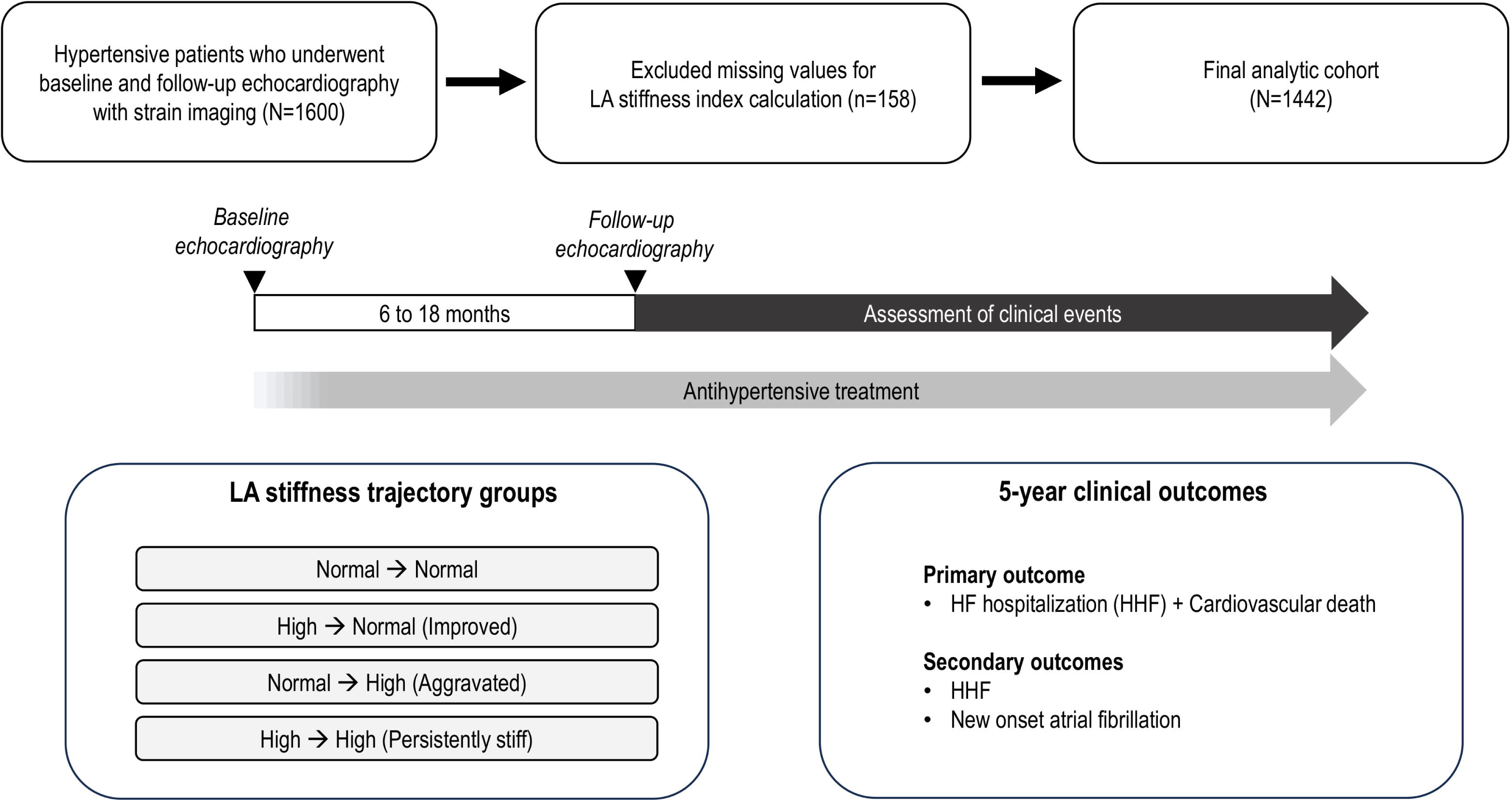
Study flow and classification of LA stiffness trajectory. Patient selection and classification into four groups based on baseline and follow-up LA stiffness index.

The study was conducted in accordance with the Declaration of Helsinki and approved by the institutional review boards of the participating centers. The requirement for informed consent was waived due to the retrospective nature of the study and minimal risk to participants.

### Echocardiographic analysis

LV dimensions and wall thickness were measured using M-mode or two-dimensional echocardiography according to current guidelines.^22^ Relative wall thickness (RWT) was calculated as twice the posterior wall thickness divided by the LV end-diastolic dimension, with values >0.42 considered increased. LV mass was estimated using the Devereux formula and indexed to body surface area to derive the LV mass index (LVMI). LA volumes were assessed using the biplane area–length method, and the LA volume index (LAVI) was calculated by indexing LA volume to body surface area. Mitral inflow velocities (E and A) and E/e′ were obtained using pulsed-wave Doppler and tissue Doppler imaging, respectively.

LV global longitudinal strain (LV-GLS) and LASr were quantified using a fully automated artificial intelligence–based software (Sonix Health Workstation, version 2.0; Ontact Health Inc., Korea).^23–26^ LV-GLS was calculated as the average peak longitudinal strain from apical 4-, 2-, and 3-chamber views, referenced to the QRS complex, and was expressed as the absolute value for ease in interpretation. LASr was calculated as the average of measurements obtained from the apical 4- and 2-chamber views, as the peak positive strain during ventricular systole. Further technical details and validation have been described previously.^25^

LA stiffness index was calculated as the ratio of E/e′ to LASr, as previously described.^15^ To evaluate longitudinal changes, patients were categorized into four groups according to temporal changes in LA stiffness index: normal–normal, improved, aggravated, and persistently stiff. A cut-off value of 0.63 derived from ROC curve analysis of follow-up measurements was applied to both time points.

### Outcomes

The primary outcome was a composite of hospitalization for heart failure (HHF) and cardiovascular (CV) death. Secondary outcomes included HHF and incident AF. For the analysis of AF, patients with a history of AF at baseline or those who developed AF during the interval between baseline and follow-up echocardiography were excluded.^19^ HHF was defined as an unplanned admission for worsening heart failure. Clinical outcomes were determined through review of medical records, structured telephone interviews, and linkage with national mortality databases.

### Statistical analysis

Continuous variables are presented as mean ± standard deviation, and categorical variables as counts and percentages. Between-group comparisons used the Student’s t-test, analysis of variance, or chi-square test as appropriate. Event-free survival was assessed using the Kaplan–Meier method and compared with the log-rank test. Cox proportional hazards regression analysis was performed to evaluate the association between LA stiffness index groups and clinical outcomes. Multivariable models were constructed by adjusting for clinically relevant covariates selected a priori. To assess the incremental prognostic value of LA stiffness index beyond clinical factors and LVMI, model performance was compared using the C-index and global chi-square statistics. A two-sided P-value <0.05 was considered statistically significant. All analyses were performed using R software (version 4.2.1; R Foundation for Statistical Computing, Vienna, Austria).

## Results

### Baseline characteristics

A total of 1,442 patients were included in the final analysis (**Table 1**). The mean age was 64.7 ± 13.1 years, and 62.2% were men. Over a 5-year follow-up period, 74 patients experienced the primary outcome. Patients who experienced the primary outcome were older and had a higher prevalence of diabetes mellitus (27.2% vs. 47.3%, p<0.001). On baseline echocardiography, they exhibited greater LA remodeling, reflected by a larger LAVI (37.5 ± 20.5 vs. 44.7 ±20.6, p=0.003), along with more impaired LV and LA strain parameters. LA stiffness index (0.50 ± 0.37 vs. 0.70 ± 0.48, p<0.001) was also higher in patients who experienced the primary outcome. Similar differences were observed in follow-up echocardiographic measurements.

**Table 1.** Baseline characteristics.

| Variables | No HHF or CV death<br>(n=1368) | HHF/CV death<br>(n=74) | P |
| --- | --- | --- | --- |
| **Clinical Factors** |  |  |  |
| Age (years) | 64.2 ± 13.0 | 73.6 ± 11.4 | <0.001 |
| Male sex | 858 (62.7%) | 39 (52.7%) | 0.108 |
| Diabetes mellitus | 372 (27.2%) | 35 (47.3%) | <0.001 |
| Dyslipidemia | 406 (29.7%) | 19 (25.7%) | 0.545 |
| Chronic kidney disease | 85 (6.2%) | 9 (12.2%) | 0.076 |
| Coronary artery disease | 259 (18.9%) | 15 (20.3%) | 0.894 |
| **Anthropometric Factors** |  |  |  |
| Body mass index (kg/m<sup>2</sup>) | 25.1±3.6 | 25.3±3.3 | 0.709 |
| SBP at baseline (mmHg) | 153.1±24.3 | 152.9±21.6 | 0.957 |
| DBP at baseline (mmHg) | 89.9±18.2 | 84.4±14.8 | 0.011 |
| Heart rate (bpm) | 71.1±20.0 | 71.1±25.5 | 0.986 |
| **Laboratory Findings** |  |  |  |
| Hemoglobin (g/dL) | 13.4±2.1 | 12.8±1.8 | 0.009 |
| Blood urea nitrogen (mg/dL) | 17.4±8.1 | 21.2±11.9 | <0.001 |
| Serum creatinine (mg/dL) | 1.0±0.6 | 1.2±0.8 | 0.021 |
| GFR (mL/min/1.73m<sup>2</sup>) | 81.0±25.4 | 68.0±25.8 | <0.001 |
| Total cholesterol (mg/dL) | 169.8±45.5 | 161.7±40.3 | 0.133 |
| **Baseline echocardiography** |  |  |  |
| LV-EDD (mm) | 48.5±6.1 | 48.9±7.1 | 0.547 |
| LV-ESD (mm) | 31.8±7.2 | 31.8±8.3 | 0.923 |
| LV-EDV (mL) | 84.3±33.8 | 81.1±35.4 | 0.445 |
| LV-ESV (mL) | 36.5±25.1 | 38.3±30.0 | 0.566 |
| LV-EF (%) | 59.6±10.2 | 57.5±12.6 | 0.089 |
| LV-MI (g/m<sup>2</sup>) | 108.2±33.4 | 114.2±31.1 | 0.130 |
| RWT | 0.43±0.10 | 0.43±0.14 | 0.667 |
| LAVI (mL/m<sup>2</sup>) | 37.5±20.5 | 44.7±20.6 | 0.003 |
| Mitral E/e' ratio | 11.9±4.8 | 13.9±4.9 | <0.001 |
| TR Vmax (m/s) | 2.3±0.5 | 2.5±0.5 | <0.001 |
| RVSP (mmHg) | 29.1±8.0 | 32.8±9.6 | <0.001 |
| LASr (%) | 29.4±11.5 | 25.8±11.1 | 0.007 |
| LV-GLS (%) | 14.7±4.6 | 12.1±4.8 | 0.006 |
| LA stiffness index | 0.50±0.37 | 0.70±0.48 | <0.001 |
| **Follow-up echocardiography** |  |  |  |
| LV-EDD (mm) | 43.4±6.1 | 44.4±7.8 | 0.777 |
| LV-ESD (mm) | 30.4±5.8 | 32.8±9.0 | <0.001 |
| LV-EDV (mL) | 80.5±33.1 | 80.7±36.2 | 0.951 |
| LV-ESV (mL) | 32.2±18.0 | 39.2±28.8 | 0.003 |
| LV-EF (%) | 61.4±8.1 | 55.4±13.8 | <0.001 |
| LV-MI (g/m<sup>2</sup>) | 106.6±31.1 | 128.4±53.9 | <0.001 |
| RWT | 0.42±0.12 | 0.45±0.15 | 0.016 |
| LAVI (mL/m<sup>2</sup>) | 36.8±15.7 | 45.3±17.9 | <0.001 |
| Mitral E/e' ratio | 11.4±4.4 | 14.3±5.1 | <0.001 |
| TR Vmax (m/s) | 2.3±0.6 | 2.4±0.6 | 0.133 |
| RVSP (mmHg) | 29.1±7.8 | 32.7±9.7 | <0.001 |
| LASr (%) | 30.6±10.4 | 26.2±11.6 | <0.001 |
| LV-GLS (%) | 16.9±4.1 | 15.4±4.7 | 0.004 |
| LA stiffness index | 0.44±0.28 | 0.69±0.42 | <0.001 |
| **Antihypertensive medication** |  |  |  |
| RAS blockers | 901 (65.9%) | 56 (75.7%) | 0.107 |
| Beta blockers | 486 (35.5%) | 25 (33.8%) | 0.857 |
| DHP-CCB | 644 (47.1%) | 39 (52.7%) | 0.410 |
| NDHP-CCB | 80 (5.8%) | 4 (5.4%) | 1.000 |
| Thiazide | 210 (15.4%) | 15 (20.3%) | 0.333 |
| MRA | 68 (5.0%) | 7 (9.5%) | 0.154 |

Values are presented as mean ± SD or n (%). P values compare patients with and without HHF and cardiovascular death.
Abbreviations: HHF, hospitalization for heart failure; SBP, systolic blood pressure; DBP, diastolic blood pressure; GFR, glomerular filtration rate; LV, left ventricular; LV-EDD, LV end-diastolic diameter; LV-ESD, LV end-systolic diameter; LV-EDV, LV end-diastolic volume; LV-ESV, LV end-systolic volume; LV-EF, LV ejection fraction; LV-MI, LV mass index; RWT, relative wall thickness; LA, left atrial; LAVI, left atrial volume index; TR Vmax, tricuspid regurgitation maximal velocity; RVSP, right ventricular systolic pressure; LARS, left atrial reservoir strain; LV-GLS, LV global longitudinal strain; RAS blockers, renin–angiotensin system blockers; DHP-CCB, dihydropyridine calcium channel blocker; NDHP-CCB, non-dihydropyridine calcium channel blocker; MRA, mineralocorticoid receptor antagonist.

### Definition of LA stiffness index trajectory

To assess the prognostic relevance of longitudinal changes in LA stiffness index, we evaluated its association with the composite outcome using both baseline and follow-up values. In standard ROC analysis, follow-up LA stiffness index demonstrated a significantly higher AUC than baseline (0.684 vs. 0.618, DeLong p=0.022, **Figure 2A**). Time-dependent AUC analysis further showed that the discriminatory performance of follow-up LA stiffness index improved progressively over time, exceeding 0.71 beyond 36 months of follow-up, whereas baseline values remained relatively stable at lower levels (**Figure 2B**). Based on these findings, trajectory groups were defined using the follow-up LA stiffness index, with a cut-off value of 0.63 applied to classify patients at both baseline and follow-up time points. Patients were subsequently categorized into four groups according to temporal changes in LA stiffness index: normal–normal (n=996), improved (n=173), aggravated (n=91), and persistently stiff (n=182) (**Figure 2C**).

**Figure 2.**
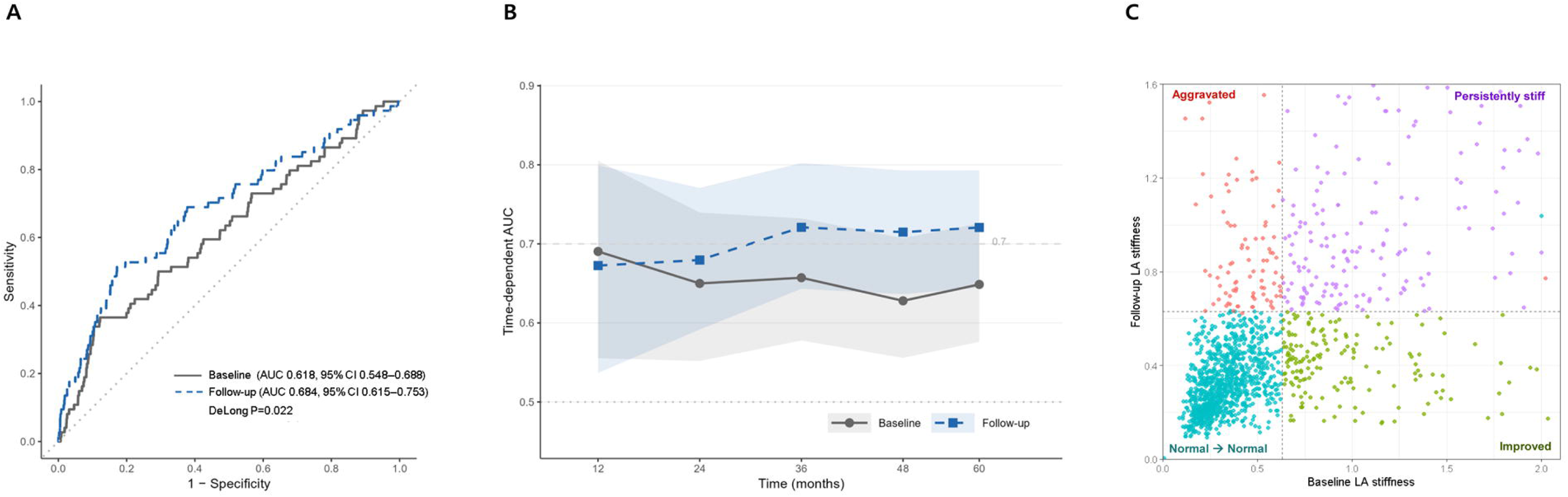
Baseline and follow-up LA stiffness index and LA stiffness index trajectory. A) ROC curves comparing the discriminatory performance of baseline and follow-up LA stiffness index. (B) Time-dependent AUC comparing baseline and follow-up LA stiffness index over the 5-year follow-up period. (C) Distribution of baseline and follow-up LA stiffness index according to trajectory group.

### Risk of the composite outcome according to LA stiffness index trajectory

Kaplan–Meier analysis demonstrated that, compared with the normal–normal group, patients in the aggravated and persistently stiff groups had a higher incidence of primary outcome (composite of HHF and CV death), whereas the improved group showed a similar risk (**Figure 3A**). These findings were consistent in multivariable Cox regression analysis. Compared with the normal–normal group, the aggravated group (adjusted hazard ratio [aHR] 2.175, 95% confidence interval [CI] 1.048–4.515, p=0.037) and the persistently stiff group (aHR 2.935, 95% CI 1.697–5.076, p<0.001) were independently associated with a higher risk of the composite outcome, while the improved group was not (aHR 0.495, 95% CI 0.151–1.617, p=0.244) (**Figure 3B**, **Table 2**).

**Figure 3.**
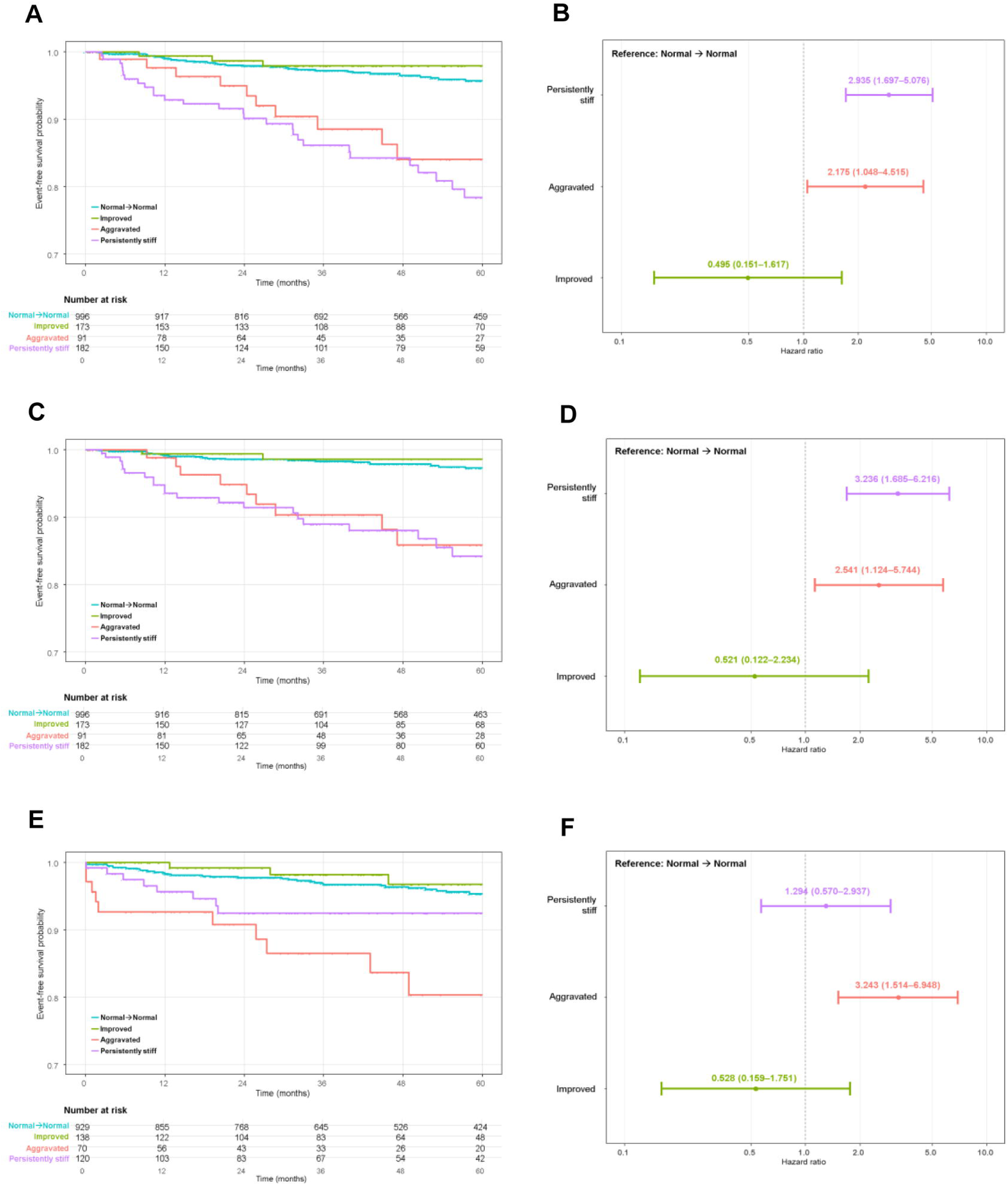
LA stiffness index trajectory and clinical outcomes. (A) Kaplan–Meier curves for the composite outcome of cardiovascular death and heart failure hospitalization. (B) Multivariable Cox regression analysis for the composite outcome. (C) Kaplan–Meier curves for heart failure hospitalization. (D) Multivariable Cox regression analysis for heart failure hospitalization. (E) Kaplan–Meier curves for new-onset atrial fibrillation. (F) Multivariable Cox regression analysis for new-onset atrial fibrillation.

**Table 2.** Multivariable analyses for clinical outcomes.

|  | Multivariate |  |  | Multivariate |  |  | Multivariate |  |  |
| --- | --- | --- | --- | --- | --- | --- | --- | --- | --- |
|  | Adjusted HR (95% CI) |  | P-value | Adjusted HR (95% CI) |  | P-value | Adjusted HR (95% CI) |  | P-value |
|  | HHF + CV death |  |  | HHF |  |  | AF |  |  |
| Age (per +1 year) | 1.060 | 1.036–1.085 | <0.001 | 1.057 | 1.029–1.085 | <0.001 | 1.037 | 1.013–1.061 | 0.002 |
| Male Sex | 1.067 | 0.664–1.715 | 0.788 | 1.000 | 0.571–1.750 | 0.999 | 1.199 | 0.684–2.101 | 0.526 |
| SBP, initial (per 1 mmHg) | 1.001 | 0.990–1.011 | 0.902 | 1.013 | 1.003–1.024 | 0.013 | 1.014 | 1.003–1.025 | 0.009 |
| DM | 1.994 | 1.254–3.172 | 0.004 | 2.494 | 1.443–4.312 | 0.001 | 1.131 | 0.641–1.996 | 0.671 |
| CKD | 1.731 | 0.855–3.506 | 0.128 | 2.589 | 1.280–5.236 | 0.008 | 1.438 | 0.609–3.395 | 0.407 |
| LV mass index, follow-up (per 1 g/m<sup>2</sup>) | 1.008 | 1.004–1.013 | <0.001 | 1.009 | 1.004–1.015 | <0.001 | 1.007 | 1.002–1.012 | 0.006 |
| LA stiffness trajectory |  |  |  |  |  |  |  |  |  |
| - Normal → Normal |  | *Referenced* |  |  | *Referenced* |  |  | *Referenced* |  |
| - Improved | 0.495 | 0.151–1.617 | 0.244 | 0.521 | 0.122–2.234 | 0.380 | 0.528 | 0.159–1.751 | 0.297 |
| - Aggravated | 2.175 | 1.048–4.515 | 0.037 | 2.541 | 1.124–5.744 | 0.025 | 3.243 | 1.514–6.948 | 0.002 |
| - Persistently stiff | 2.935 | 1.697–5.076 | <0.001 | 3.236 | 1.685–6.216 | <0.001 | 1.294 | 0.570–2.937 | 0.538 |
| C-index | 0.776 |  |  | 0.833 |  |  | 0.732 |  |  |

Hazard ratios (HRs) are shown with 95% confidence intervals (CIs).
Abbreviations: HHF, heart failure hospitalization; CV, cardiovascular; SBP, systolic blood pressure; DM, diabetes mellitus; CKD, chronic kidney disease; LA, left atrial.

### Association between LA stiffness index trajectory and secondary clinical outcomes

To further evaluate the prognostic relevance of LA stiffness index trajectory, we analyzed secondary clinical outcomes, including HHF, as well as incident AF (**Figure 3**). For HHF, Kaplan–Meier analysis showed a pattern similar to that observed for the composite outcome, with higher event rates in the aggravated and persistently stiff groups than in the normal–normal group, while the improved group showed a comparable risk (**Figure 3C**). In multivariable Cox regression analysis, both the aggravated group (aHR 2.541, 95% CI 1.124–5.744, p=0.025) and the persistently stiff group (aHR 3.236, 95% CI 1.685–6.216, p<0.001) remained independently associated with a higher risk of HHF, using the normal–normal group as the reference (**Figure 3D**, **Table 2**).

For incident AF, a total of 1,255 patients without a history of AF at baseline and who did not develop AF during the interval between baseline and follow-up echocardiography were included in the analysis. Over a median follow-up of 51.1 months (interquartile range [IQR] 27.9–60.0), the median number of electrocardiographic evaluations per patient was 8 (IQR 4–15). Kaplan–Meier analysis showed a higher incidence of new-onset AF in the aggravated group than in the normal–normal group, while the improved group had a similar risk (**Figure 3E**). This pattern was supported by multivariable Cox regression analysis, in which the aggravated group was independently associated with a higher risk of AF compared with the normal–normal group (aHR 3.243, 95% CI 1.514–6.948, p=0.002) (**Figure 3F**, **Table 2**). By contrast, the improved group was not associated with an increased risk of AF (aHR 0.528, 95% CI 0.159–1.751, p=0.297). Although the persistently stiff group showed a numerically higher incidence of new-onset AF, this association was not statistically significant after multivariable adjustment (aHR 1.294, 95% CI 0.570–2.937, p=0.538).

### Incremental prognostic value of LA stiffness index beyond clinical factors and LVMI

To evaluate the incremental prognostic value of LA stiffness index beyond clinical factors and LVMI, model performance was assessed with sequential addition of variables (Figure 4). Addition of LVMI to the clinical model improved model fit (Δχ²=19.3, p<0.001; C-index 0.760). When entered as continuous variables, LA stiffness index provided the greatest incremental value (Δχ²=12.6, p<0.001; C-index 0.768), whereas LASr (Δχ²=4.5, p=0.034) and E/e′ (Δχ²=3.8, p=0.052) showed modest or non-significant incremental value (**Figure 4A**, **4B**). When dichotomized using Youden index-derived cut-offs (LA stiffness index ≥0.63, E/e′ ≥12.6, LASr <21.1%), all three parameters demonstrated significant incremental value, with LA stiffness index showing the greatest chi-square (Δχ²=15.9, p<0.001; C-index 0.772), followed by E/e′ (Δχ²=10.5, p=0.001) and LASr (Δχ²=11.0, p<0.001). Incorporation of LA stiffness index trajectory further increased the incremental chi-square to 20.1 (p<0.001; C-index 0.776) (**Figure 4C**, **4D)**

**Figure 4.**
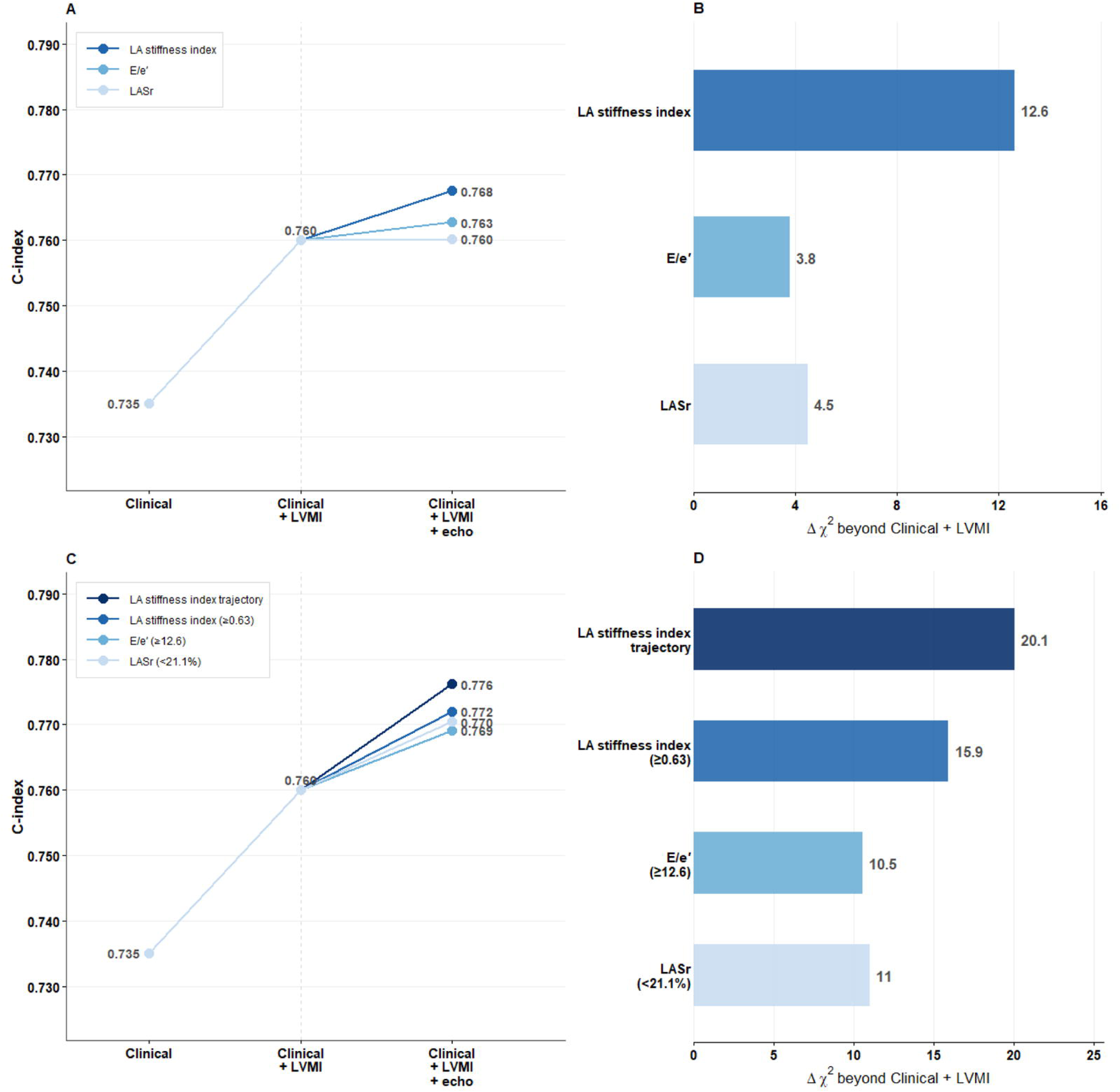
Incremental prognostic value of LA stiffness index beyond clinical variables and LV mass index. (A) C-index with sequential addition of LA functional parameters as continuous variables. (B) Incremental chi-square (Δχ²) of LA functional parameters as continuous variables beyond Clinical + LVMI. (C) C-index with sequential addition of LA functional parameters as categorical variables. (D) Incremental chi-square (Δχ²) of LA functional parameters as categorical variables beyond Clinical + LVMI.

## Discussion

In this study, we evaluated the association of LA stiffness index with clinical outcomes in patients with hypertension. The main findings are as follows. First, a higher LA stiffness index was associated with an increased risk of the composite outcome of HHF and CV death, with consistent associations observed at both baseline and follow-up measurements. Second, longitudinal changes in LA stiffness index provided additional prognostic information, as patients with improvement showed a risk comparable to those with persistently normal values, whereas those with aggravated values had a higher risk of the composite outcome comparable to those with persistently elevated LA stiffness index. Third, LA stiffness index, especially its longitudinal change, demonstrated incremental prognostic value beyond LVMI, supporting its independent clinical relevance. Taken together, these findings suggest that both baseline LA stiffness index and its longitudinal change are clinically meaningful markers for risk stratification in hypertensive patients.

### Left atrial remodeling in hypertension: beyond left ventricular hypertrophy

Hypertension induces structural and functional remodeling of the LV, which has been a central focus in the assessment of target organ damage.^27^ Chronic pressure overload leads to LVH, myocardial fibrosis, and impaired ventricular function, ultimately contributing to adverse cardiovascular outcomes.^28^ Further, in patients with hypertension, the longitudinal change in the LV remodeling (i.e., development or regression of LVH) has been suggested as a key determinant of prognosis.^17,29^ However, compared to the detailed recommendations and the wide utilization of LV remodeling in patients with hypertension, the structural and functional consequences of LA have received relatively less attention in the context of hypertension, despite its close physiological coupling with the LV. Elevated LV filling pressure, which accompanies hypertensive remodeling, is transmitted to the LA and leads to progressive LA remodeling. As a result, the LA reflects the cumulative impact of chronic hemodynamic burden, incorporating both structural and functional alterations over time.^30,31^ Therefore, assessment of LA remodeling may provide additional insights into disease progression beyond conventional LV-focused parameters.

### LA stiffness index reflects both LV filling pressure and LA functional adaptation

Several previous studies have investigated the hemodynamic impact of hypertension on LA structure and function, demonstrating significant associations between hypertension and LA remodeling.^4,5^ However, conventional echocardiographic parameters such as E/e′, LAVI, and tricuspid regurgitation velocity have limited sensitivity for capturing intrinsic LA function under varying loading conditions and limited specificity that necessitates integrated interpretation of multiple parameters.^15,32,33^

More recently, LA strain has emerged as a sensitive marker of atrial dysfunction and reflects intrinsic myocardial properties;^13,34^ however, it is influenced by loading conditions and may not fully capture structural remodeling such as fibrosis.^12,15^ In this context, the LA stiffness index, derived from E/e′ and LASr, provides a composite measure that integrates both LA function and LV filling pressure. Conceptually, stiffness is defined as the ratio of pressure change to volume change (ΔP/ΔV). E/e′ is a surrogate of LV filling pressure, whereas LASr reflects atrial deformation as a surrogate of volume change. Combining these two components, LA stiffness index approximates the LA pressure-volume relationship, with higher values indicating elevated LA pressure and reduced reservoir deformation.^15,16,35^ Consistent with this, LA stiffness index demonstrated incremental prognostic value beyond LVMI and clinical variables in the present study, outperforming LASr and E/e′ individually, suggesting that the integration of these two components captures prognostically relevant information that neither parameter alone can provide.

### Prognostic significance of LA stiffness index trajectory

One of the key strengths of the present study is the incorporation of serial echocardiographic assessments, which enabled evaluation of longitudinal changes in LA stiffness index. While both baseline and follow-up LA stiffness indices were associated with clinical outcomes, trajectory-based analysis provided prognostic information beyond a single time-point measurement, underscoring the clinical value of serial echocardiographic evaluation in hypertensive patients. Patients in the improved group showed a risk comparable to those with persistently normal values, whereas those in the aggravated group exhibited a significantly higher risk of the composite outcome. These findings suggest that improvement in LA stiffness index may reflect favorable structural and functional remodeling associated with better clinical outcomes, while worsening of LA stiffness index may indicate ongoing adverse remodeling despite treatment. In our previous work, reductions in systolic blood pressure were associated with regression of left ventricular hypertrophy, as reflected by changes in LV mass index.^17^ In the present study, changes in LA stiffness index were significantly correlated with both blood pressure reduction (r=0.13, P<0.001) and changes in LV mass index (r=0.25, P<0.001, **Supplementary Figure 1**), suggesting that antihypertensive treatment influences LA remodeling through parallel changes in blood pressure and LV structure. These findings highlight the close interplay between LV and LA remodeling and underscore the clinical value of follow-up echocardiographic assessment in capturing these longitudinal changes.

Regarding incident AF, the aggravated group demonstrated a significantly higher risk, whereas the persistently stiff group did not reach statistical significance despite a numerically elevated hazard ratio. This discrepancy may partly reflect the exclusion of patients with prevalent or interval-onset AF prior to the follow-up echocardiogram, which disproportionately affected the persistently stiff group given their chronically elevated LA burden. Additionally, AF development is influenced by a broader range of factors beyond LA stiffness, such as age, autonomic remodeling, systemic inflammation, and comorbidities such as obesity.^36–38^

### Clinical implications

In patients with hypertension, echocardiographic evaluation remains essential for assessing target organ damage and for risk stratification. While conventional parameters such as LVH and LA enlargement have been widely used, emerging evidence suggests that more advanced functional indices using echocardiography may provide additional insights into cardiac remodeling and prognosis. In particular, the LA stiffness index, which reflects not only the innate LA function but also its relationship with LV diastolic function and filling pressure, may have potential as an indicator of clinical course and treatment response in patients with hypertension. Furthermore, the findings of this study highlight the importance of considering longitudinal changes in these indices, rather than relying solely on single time-point measurements, for risk stratification. These findings support the clinical utility of incorporating both conventional and advanced echocardiographic parameters, as well as their temporal changes, in the comprehensive evaluation of hypertensive heart disease.

### Limitations

Several limitations of this study should be acknowledged. First, the retrospective design of the STRATS-HHD registry introduces the potential for selection bias, and causal inferences cannot be drawn from the observed associations. Second, the cut-off value for LA stiffness index used to define trajectory groups was derived from follow-up measurements within the same cohort and was not validated in an external population, which may limit the generalizability of the trajectory classification. Third, although antihypertensive medications were recorded at baseline, their specific types and changes over the follow-up period were not incorporated into the multivariable models, and therefore the influence of treatment-specific effects on LA remodeling cannot be fully assessed. Fourth, the number of primary outcome events was relatively small, which may have limited the statistical power for subgroup analyses, particularly for the improved group where confidence intervals were wide. Fifth, the study population consisted of relatively high-risk patients from two tertiary referral centers in Korea. Therefore, our findings may not be directly generalizable to other ethnic groups or healthcare settings.

### Conclusion

In patients with hypertension, LA stiffness index, a composite measure integrating LA function and LV filling pressure, was associated with adverse clinical outcomes and provided prognostic information beyond LV-focused assessment. Serial changes in LA stiffness index further refined risk stratification, with improvement indicating a favorable clinical course and aggravation identifying patients at higher risk.

### Perspectives

The present findings highlight the potential of serial LA stiffness index assessment as a practical tool for monitoring cardiac remodeling and refining risk stratification in patients with hypertension. As hypertensive heart disease represents a continuum from subclinical remodeling to overt heart failure, integrating both LV and LA longitudinal assessment may provide a more comprehensive framework for identifying high-risk patients and guiding individualized treatment strategies. Further prospective studies in diverse populations are warranted to validate the clinical utility of LA stiffness index trajectory in hypertensive heart disease.

## Novelty and Relevance

### What Is New?

- LA stiffness index was evaluated as a prognostic marker in a large hypertensive cohort, demonstrating independent associations with adverse cardiovascular outcomes beyond conventional LV-focused parameters.
- Longitudinal changes in LA stiffness index provided additional prognostic information beyond single time-point measurement, with trajectory-based classification demonstrating greater incremental prognostic value than individual LA functional parameters including LASr and E/e′.

### What Is Relevant?

- LA stiffness index, integrating LA function and LV filling pressure, extends hypertensive heart disease assessment beyond conventional LV-focused parameters, and its longitudinal changes provide additional prognostic information in hypertensive patients.

### Clinical/Pathophysiological Implications?

- Evaluation of both LV and LA remodeling may improve comprehensive risk stratification in hypertensive heart disease.
- Serial assessment of LA stiffness index may serve as a practical marker of treatment response and disease progression following antihypertensive therapy.

## Data Availability

All data produced in the present study are available upon reasonable request to the corresponding authors.

## Abbreviations

LVH: left ventricular hypertrophy
LV: left ventricular
LA: left atrial
HF: heart failure
AF: atrial fibrillation
STRATS-HHD: Strain for Risk Assessment and Therapeutic Strategies in Patients With Hypertensive Heart Disease
RWT: relative wall thickness
LVMI: left ventricular mass index
LAVI: left atrial volume index
LASr: left atrial reservoir strain
LV-GLS: left ventricular global longitudinal strain
ROC: receiver operating characteristic
HHF: hospitalization for heart failure
CV: cardiovascular
AUC: area under the curve
aHR: adjusted hazard ratio
CI: confidence interval
IQR: interquartile range
ΔP/ΔV: change in pressure to change in volume

## Acknowledgements

None.

## Data availability statement

The datasets used and analyzed during the current study are available from the corresponding author on reasonable request.

---

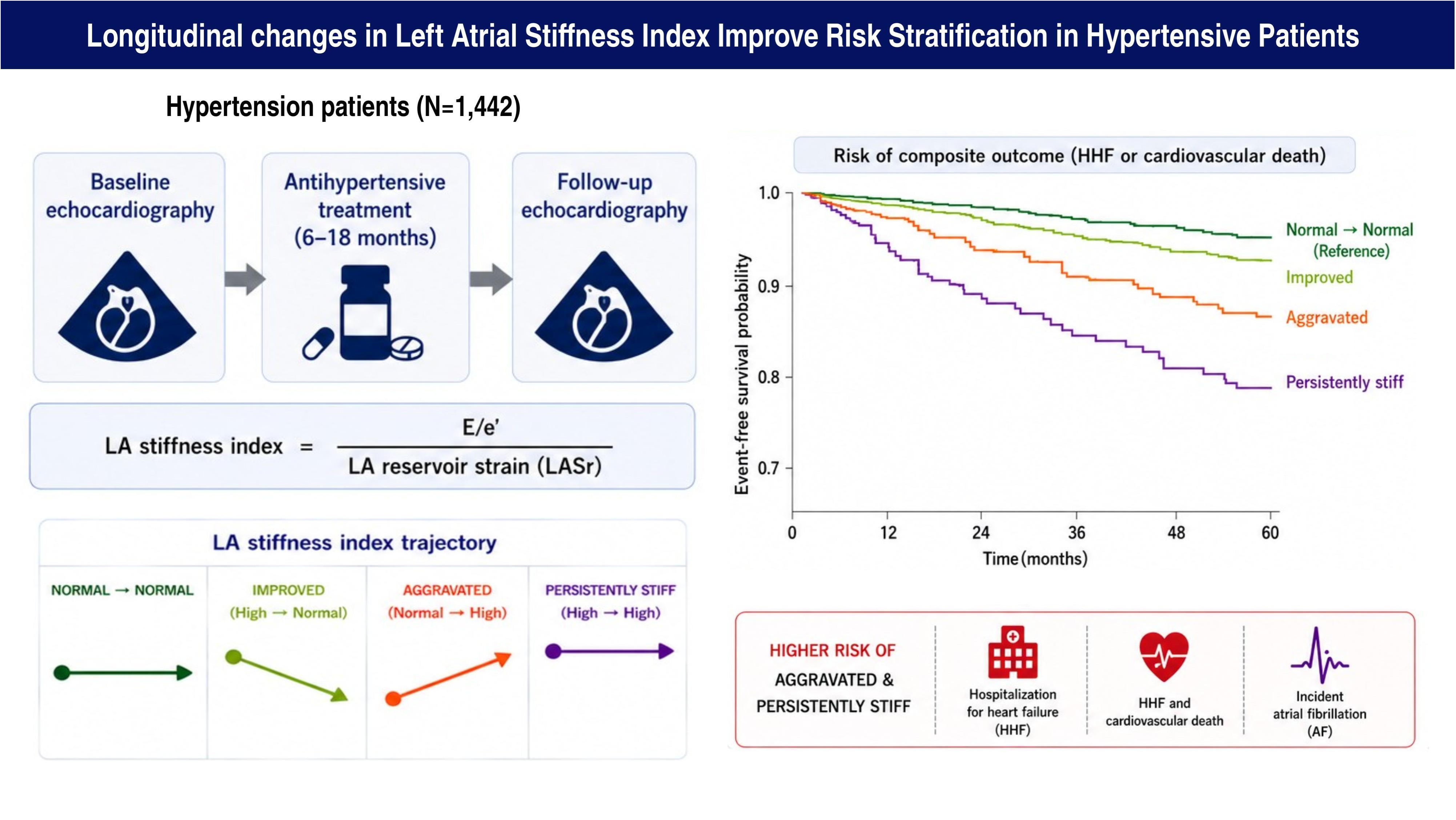

