## Supplementary Figure for "Longitudinal Changes in Left Atrial Stiffness Index Improve Risk Stratification in Patients with Hypertension"

**Supplementary Figure 1. Correlations between changes in LA stiffness index and changes in blood pressure and LV structural parameters.**

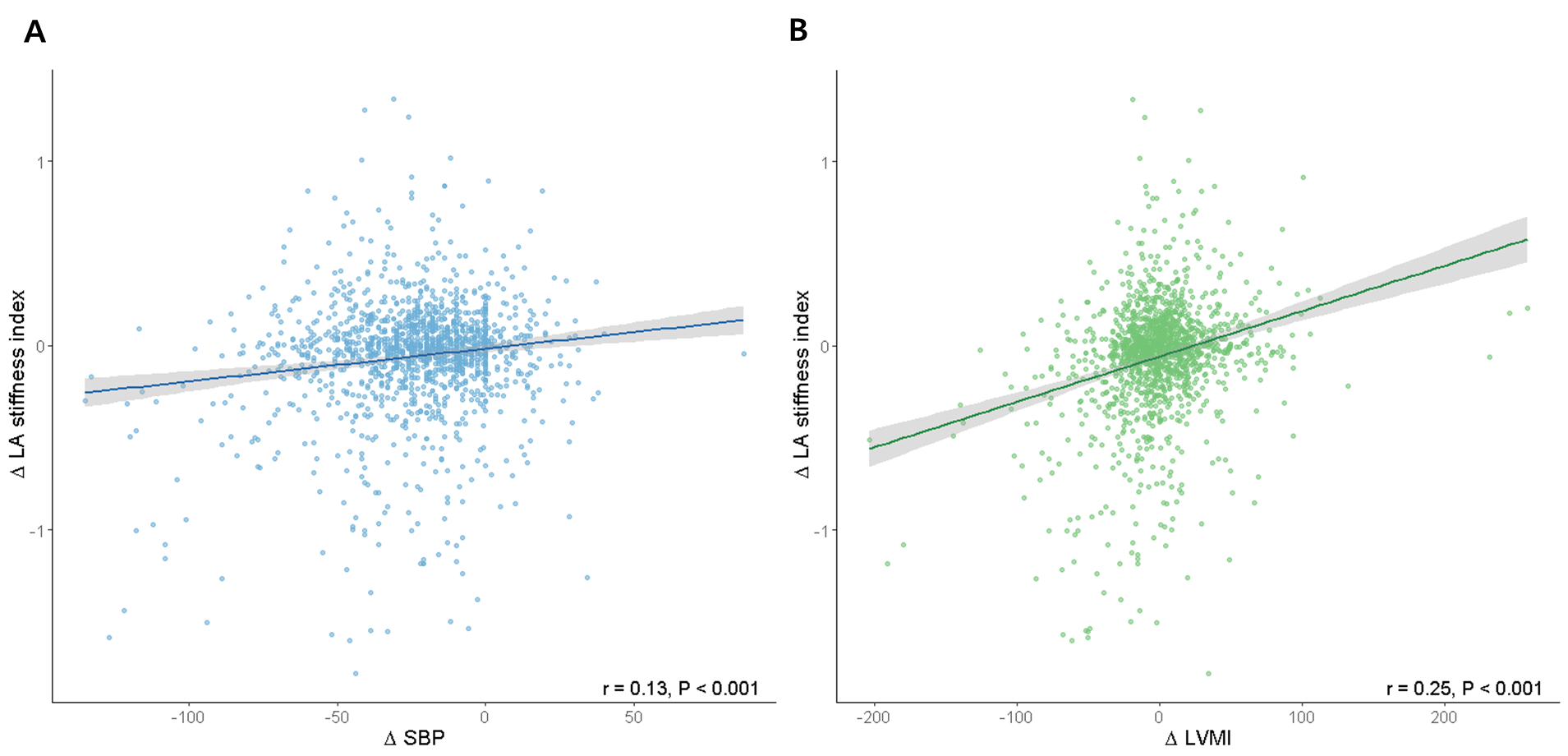

(A) Correlation between change in systolic blood pressure (ΔSBP) and change in LA stiffness index.

(B) Correlation between change in LV mass index (ΔLVMI) and change in LA stiffness index.
